# Data analysis of COVID-19 wave peaks in relation to latitude and temperature for multiple nations

**DOI:** 10.1101/2021.08.12.21261974

**Authors:** Mayuri Jain, Sukhada Aloni, Pravin Adivarekar

**Affiliations:** A. P. Shah Institute of Technology, Thane, Maharashtra, India 400615

**Keywords:** COVID-19, Temperature, latitude

## Abstract

It was observed that the multiple peaks of coronavirus disease-19 (COVID-19) appeared in different seasons in different countries. There were countries where the COVID-19 peak occurred during extremely low temperatures, such as Norway, Canada and on the other hand there were countries with high-temperature ranges such as Brazil, India, UAE. Most of the high-latitude countries received their outbreak in winter and most of the countries near the equator mark the outbreak during the summer. Most of the biological organisms have their growth dependant on the temperature, and hence we explored that if there is any relation of temperature versus COVID-19 outbreak in the particular country. It was also seen that people are not behaving differently during the peak of the COVID-19 wave, hence it was important to know whether the COVID-19 virus has evolved or the global temperature variation caused these multiple peaks. This work focuses on finding the effect of temperature variation on the COVID-19 outbreak. We used Levenberg–Marquardt technique to find the correlation between the temperature at which COVID-19 outbreak peaks and the latitude of the particular country. We found that between the temperature range of 14 *°*C to 20 *°*C spread of the COVID-19 is minimal. Based on our results we can also say that the COVID-19 outbreak is seen in lower temperature (0 *°*C to 13 *°*C) ranges as well as in the higher temperature ranges (21 *°*C to 35 *°*C). The current data analysis will help the authorities to manage their resources in advance to prepare for any further outbreaks that might occur in the COVID-19 or even in the next pandemic.

## 1 Introduction

The coronavirus pandemic has affected most of the world, it needs the highest degree of data analysis to avoid any further catastrophes [5]. The growth of any biological species is temperature dependant and the spread can be controlled with the increase and decrease of temperatures [18]. There are many types of research related to viruses, showing that there is indeed a correlation between temperature and humidity and infection due to a particular virus [7, 17]. Most of the time the increase in temperature causes a reduction in the infection spread. We tried to address the question about COVID-19 infection dependence on temperature. Recently published studies found mixed results about the COVID-19 infection spread and its temperature dependence. It was also proved that virus transmission gets affected with humidity above 6gm/kg and also seasonal variations.

Corona viruses are Ribonucleic Acid (RNA) type viruses that have been studied in detail for both medical and veterinary importance. It was well established that coronavirus can be killed by ultraviolet (UV) rays, extreme pH, and also changing physical and chemical conditions. The outer structural protein of coronavirus becomes inactive at certain temperature ranges. This particular temperature dependence where the spread of the virus can be seen differently at various ranges of latitude. The climate cannot be the only reason to affect the COVID-19 transmission. There are other variables such as social distancing, age, GDP, health conditions (Diabetes, Coronary heart diseases), physical activity, medical infrastructure, etc. For this particular manuscript, we only investigate the effect of latitude and temperature on the peaks of the COVID-19 outbreak for selected 12 countries. We have developed an algorithm based on the sum of the sine method to predict the temperature at which the COVID-19 peak will occur with respect to latitude. We validated our prediction algorithm with blinded studies using 7 more countries.

Figure 1 shows the proposed system workflow. Covid-19 counts, temperature range, and latitude were collected from different data sources. Based on the data obtained and Levenberg–Marquardt algorithm Sum of the Sine equation first order fitting were obtained. The output analysis consisted of Latitude versus Covid peak, Temperature versus Covid count, and Histogram of Number of Countries peaking at specific Temperature. In the present study, we have collected data about COVID-19 cases reported per day in 12 countries of the world. To find the relation between temperature and COVID-19 case peaks we also collected one-year temperature data. After carefully analyzing the data we could establish that COVID-19 cases are not spiking in the specific temperature range of 14 *°*C to 20 *°*C. We could also establish the relationship between latitude and corresponding temperature at which the COVID-19 outbreak peak can occur. To validate our results with 7 countries chosen at random which were not part of the training dataset of 12 countries and could find that accuracy of our prediction is 81.1 %

**Fig. 1.**
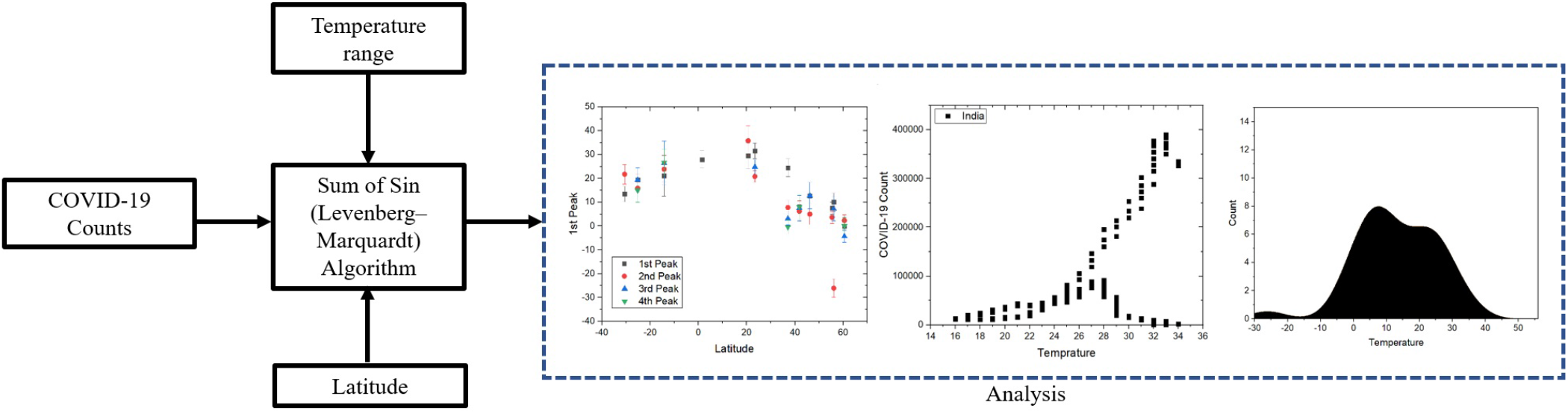
Workflow for the proposed system. Counts of COVID-19, temperature range, and latitude were gathered from several data sources. Sum of Sine equation with first-order fits well to the data obtained with help of Levenberg–Marquardt technique. The data analysis included a peak in the COVID-19 count versus the latitude of the country, COVID-19 cases plotted against temperature, and a histogram of the number of countries peaking at certain temperatures.

## 2 Literature review

Notari [11] showed that there exists a definite temperature correlation with the exponential growth of covid. The work claims that there is about a 40 % reduction in the spread of COVID cases with a temperature of about 25 *°*C compared to 5 *°*C. However, the work could not fit the majority of warmer and poorer countries, Author claimed the reason could be less intense testing. Smit *et al*. [15] investigated multiple published papers on COVID-19 and its spread with respect to environmental factors such as Temperature and Humidity. The authors claim that there could be misdiagnosis between COVID-19 and seasonal flu or influenza. They did not find any strong correlation between environmental factors and COVID growth, contradicting many other claims. Briz *et al*. [1] described hot weather may not be a strong candidate in preventing the spread of COVID-19. The work suggested that there is the general assumption that weather patterns may affect the transmission and spread of the viral droplets that cause the infection. Runkle *et al*. [14] studied the effect of temperature and humidity for the short term on COVID-19 infection. They found that there exists definite relation between humidity and COVID-19 spread. They used a 7-days distributed lag nonlinear model. Marvi *et al*. [10] claimed that, there is a slow rate of infection spread for regions with extreme temperatures. They have also considered population density, number of tests conducted per day, number of days from the first infection.

Goyal *et al*. [8] studied the effect of temperature on the spread of COVID-19. They also found that the colder states have an inverse parabolic relation with respect to the spread of COVID i.e, as temperature decreases the cases were increasing. They found that hotter states show a consistent increase in cases and are of exponential nature and hence concluded that temperature does not appear as a significant determinant of COVID-19 spread.

Demongeot *et al*. [6] found that temperature reduces the spread of new COVID cases. They found that the velocity of COVID-19 spread was temperature-dependent. They use 21 countries’ data and analyzed using ARIMA(Auto Regressive Integrated Moving Average).

Byun *et al*. [3] studied data for a complete one year to find seasonal factors affecting COVID transmissibility. They have studied 50 countries and found that most of the countries have an inverse correlation with COVID transmission. They mentioned that COVID transmission will decrease with temperature, absolute humidity, and solar radiation and found that COVID infection spreads with latitude and precipitation. Burra *et al*. [2] tried to correlate latitude and temperature with COVID-19 spread. They found a strong correlation between temperature and mortality. They also found some relation between latitude and COVID-19 spread, however longitude does not matter. One of their major achievement was gnome sequence is completely uncorrelated with temperature and geographic coordinates. Ren *et al*. [12] found that temperature increase was the main reason for the reduction in COVID-19 cases in China. The geographical locations without central heating and temperature control have a strong effect in restricting COVID-19 spread. They have only studied data from one country i,e China and a temperature range from 4 *°*C. They applied an analysis of variance(ANOVA) statistical model. Juni *et al*. [9] found that temperature and latitude do not have any association with COVID-19 spread. They considered 144 geolocations in Australia, Canada, and the United States. They specifically excluded China, Italy, Iran, and South Korea. They also stated that hotter weather has no association with COVID spread. Finally, they concluded both latitude and temperature could play a role in a pandemic however they mention that they got opposite results. According to them the summer is not going to end the pandemic.

## 3 Methodology

### 3.1 Equipment and Softwares

A computer with an Intel Core i7 6700K processor and 24 GB DDR4 ram was used to do all the computations. MATLAB software version 2019a was used for all the computations and data fitting.

### 3.2 Data collection

The COVID patient count data was acquired from “Our-World” database from 20 February 2020 till 18 May 2021 [13]. The database consists of 207 country profiles including 195 countries and different continents and their combinations, out of which we have selected 12 countries. These 12 countries were India, Italy, France, Brazil, South Africa, Canada, Singapore, UK, UAE, US, Norway, Australia. Figure 2 shows daily new confirmed cases (7-Day Average) plotted in log format obtained from dataset. The X-axis shows Time (from 20th February 2020 till 18th May 2021). The Y-axis shows the number of confirmed COVID-19 cases in logarithmic form. Total 12 countries were considered based on their position on Earth (Latitude) and multiple peaks of the COVID-19 wave. India (Greenish Blue) is the highest with the maximum number of cases and Australia (Dark Green) is the lowest. The choice of countries was done on basis of latitude variation, population diversity, and countries having varying peaks (e.g. from a country with a single COVID-19 outbreak peak to countries with 4 peaks in a year). The temperature data was collected from the weather-atlas and climate-data datasets [4]. The temperature reading consists of roughly 1.5-year data starting from 1st January 2020 onwards till 18th May 2021. The values include the daily average temperature of a country, maximum temperature, and minimum temperature (fig. 3b). The latitude information was collected from Simplemaps dataset [16].

**Fig. 2.**
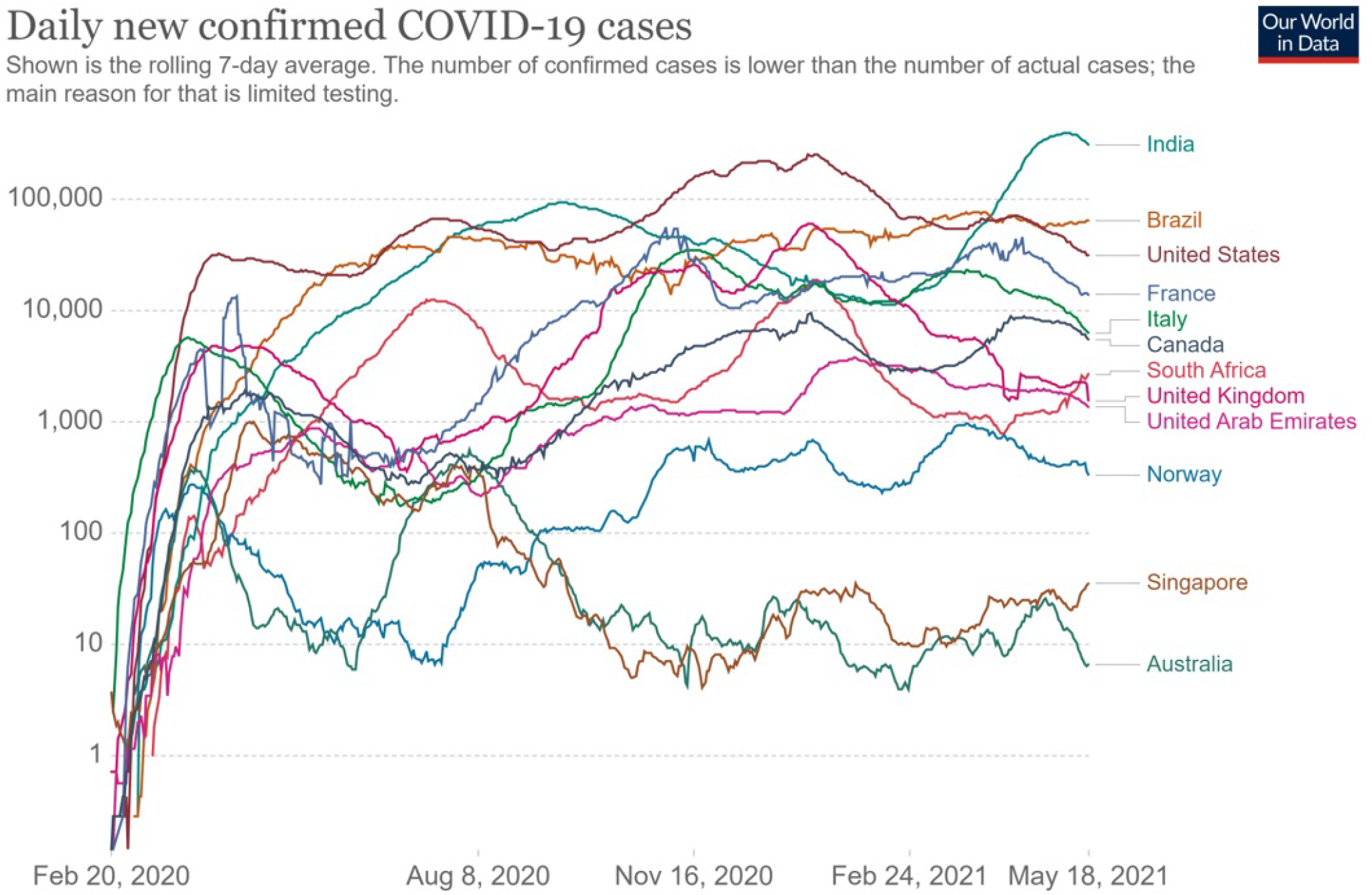
The number of new confirmed cases per day (7-day average) is plotted in semi-log format. On the X-axis, time is shown for 453 days, from February 20, 2020, to May 18, 2021. The number of confirmed COVID-19 cases is shown in the logarithmic form on the Y-axis. A total of 12 countries were considered based on their geographic location (latitude) and multiple peaks of the COVID-19 wave. India (Greenish Blue) has the highest number of cases, while Australia (Dark Green) has the lowest.

**Fig. 3.**
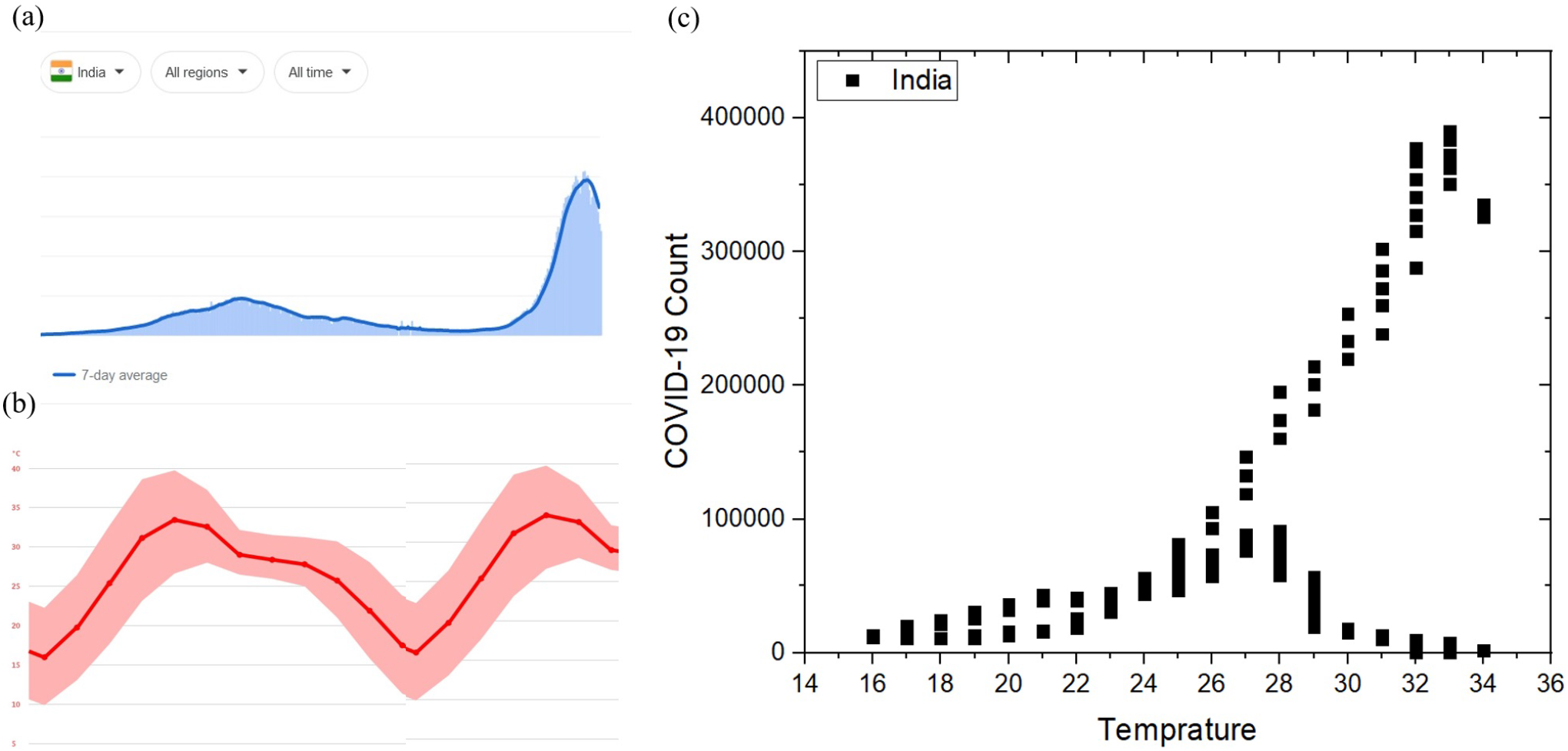
Effect of temperature on COVID-19 count in India. (a) COVID-19 distribution curve plotted with 7 days average value. There can be two clear peaks seen. The count was recorded from 20 February 2020 till 18 May 2021. (b) Average temperature distribution of India for the period of 1 January 2020 till 18 May 2021 the break in the gray lines indicates year-end. (c) The plot of temperature vs COVID-19 counts for each day. It was observed that temperatures below 21 *°*C do not have a significant COVID count. Temperature around 27 to 35 *°*C seen a totally different kind of distribution, one peaking towards 4 Lakh COVID-19 count per day and other tending to zero. These two separate behaviors were observed during second and first wave peaks respectively.

### 3.3 Peak assessment method

The data for each day was carefully analyzed and the slope (s) was computed between consecutive days using equation 1. once the slope changes the direction from positive to negative and if the magnitude at one day before slope change is at least 50 % more than the average value of last 30 days, then one day before that day is considered as a day at which peak occurred.

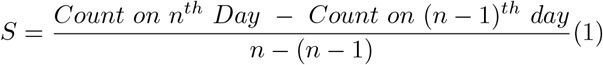

For the peaks which are very close by i.e. within 75 % of the peak of the bell-shaped curve the maximum magnitude was considered and all the nearby values were neglected avoiding over-fitting of the data.Two such mapped peaks are shown in Fig. 4c and Fig. 4d

**Fig. 4.**
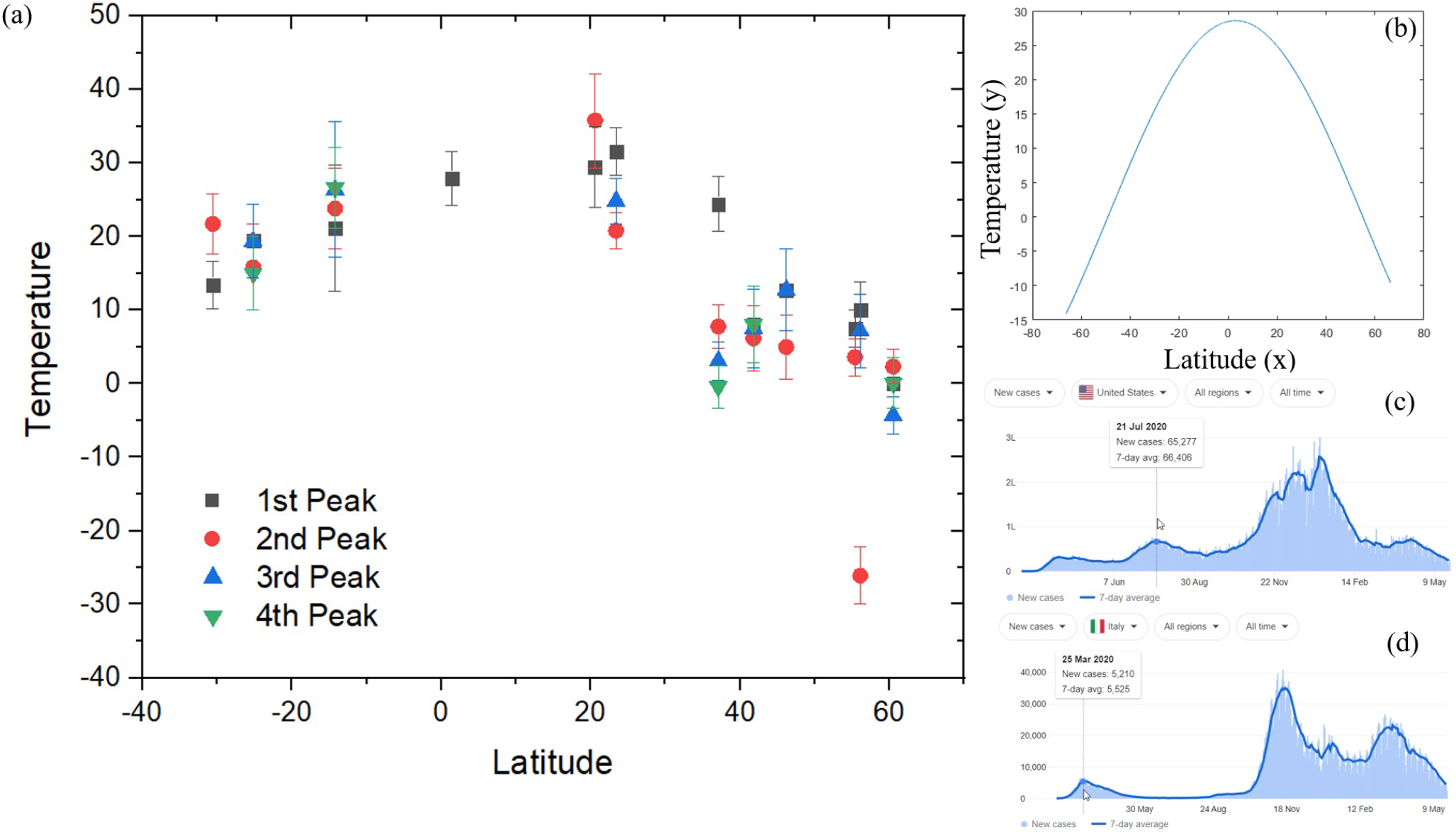
(a) A plot of latitude versus temperature for 12 nations throughout the world. Temperature at which COVID-19 peak occur steadily decreases, when latitude increases to the south or north. In the majority of cases, the temperature with the second, third, and fourth peaks were found to be at a greater temperature than the initial peak. Singapore is the only country out of the 12 deemed to have only a single peak, while Canada is the only country with a COVID-19 peak occurring at a temperature of −30 °C. (b) A plot of latitude versus temperature produced from the best fitting to the data points in (a) using the Levenberg–Marquardt algorithm and the first-order Sum of Sine equation. (c) COVID-19 count for the United States shows that the second peak occurred on July 21, 2020, with a seven-day average of 66406 cases. (d) The first peak of COVID-19 cases in Italy occurred on March 25, 2020, with a seven-day average of 5525 cases.

### 3.4 Data fitting

Once the data for each countries peak was extracted, then we found the range of temperature on the day at which the peak occurred. The plot of all the peaks versus latitude was computed as shown in Fig. 4 (a). For data fitting, curve fitting code was written in MATLAB. After multiple attempts of fitting and systematic studies of all the fitting, we found the Levenberg–Marquardt algorithm and the first-order Sum of the Sine equation gives a maximum value of goodness of fitting (i.e. Coefficient of determination *R*^2^ almost equal to 1). The fitting obtained was given in equation 2

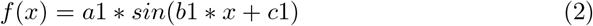

Coefficients (with 95 % confidence bounds) are

a1 = 28.57 and (min = 27.09, max = 30.06)

b1 = 0.03015 and (min = 0.02891, max = 0.03138)

c1 = 1.482 and (min = 1.429, max = 1.535)

## 4 Results and Discussion

Figure 3 shows the effect of temperature on COVID-19 peaks and cases. (a) The effect of temperature on COVID-19 cases was found to be divided into two regions, namely, Low-temperature regions indicated in light blue color and high-temperature regions indicated in light pink color. The total number of COVID-19 cases in the Low-temperature region (0 *°*C to 13 *°*C) was found to be comparable with the high-temperature region (21 *°*C to 35 *°*C). (b) Histogram of the number of countries peaking at specific temperatures. Roughly 8 countries saw the peak of their COVID-19 wave at approximately 7 *°* C. this bi-modal distribution has a second peak at around 25 *°*C where roughly 7 countries were affected.

Figure 4 shows that temperature affects the COVID-19 count for India. COVID-19 distribution curve with the 7-day average value displayed in Fig. 4a. There may be two distinct peaks to the wave. The count was taken from 20 February 2020 to 18 May 2021. Figure 4b shows the Indian average temperature distribution from 1 January 2020 to 18 May 2021; the break in the gray lines denotes the end of the year. For each day, the plot of temperature versus COVID-19 count is shown in Fig. 4c. Temperatures below 21 degrees were found to have no significant COVID count. Temperatures ranging from 27 to 35 degrees produced a completely distinct distribution, with one trending towards 4 lakh COVID-19 cases each day and the other trending towards nil.

Figure 5 (a) shows the plot of latitude versus temperature for 12 different countries in the world. As the latitude increases towards the South or North the COVID-19 peaks decrease gradually. In most of the cases out of four peaks considered the temperature with second, third, fourth peaks were found to be higher than the first peak. Singapore is the only country out of the considered 12 to have only a single peak and Canada is the only country that has a COVID-19 peak occurring in the freezing temperature at −30 *°*C. Fig. 5 (b) shows the Latitude versus temperature plot derived from best fitting to the data points in fig. 5 (a) using Levenberg–Marquardt algorithm with the help of the first-order Sum of the Sine equation. Fig. 5 (c) shows COVID-19 count for the United States indicating the second peak occurred on 21 July 2020 with seven day average of 66406 cases. Fig. 5 (d) shows COVID-19 cases for Italy’s first peak occurred 25 March 2020 with seven-day average cases of 5525.

**Fig. 5.**
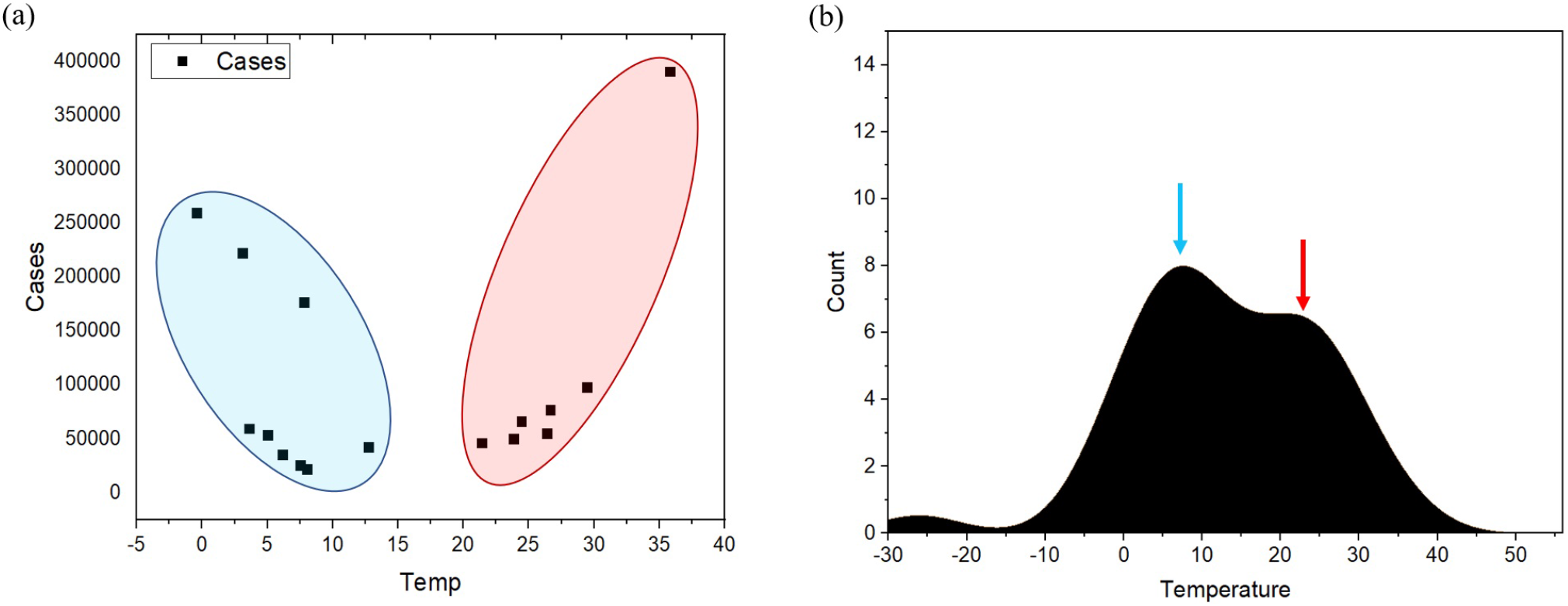
Effects of temperature on COVID-19 peaks and cases. (a) The temperature influence on COVID-19 cases was discovered to be divided into two zones, namely low-temperature zone indicated in light blue color and high-temperature zone indicated in light pink color. The overall number of COVID-19 cases detected in the low-temperature zone (0 *°*C to 13 *°*C) was comparable to the high-temperature zone (21 *°*C to 35 *°*C). (b) Histogram showing the number of countries (Frequency on Y-axis) where COVID-19 peak occurs at a specific temperature (X-axis). Approximately 8 countries experienced the peak of their COVID-19 wave at around 7 *°*C. This bimodal distribution has a second peak at about 25 *°*C, affecting around 7 countries.

As shown in Table 1, we tested 7 different countries which were not part of the training procedure. For all the blind testing we chose latitude as the input parameter and found out the temperature at which the COVID outbreak peak will occur. For suitable distribution, we chose lower values of latitude such as Indonesia with a latitude of −0.8 and chose the countries with higher latitude values such as France and New Zealand. During the testing, we ensured both positive and negative latitude were covered equally. For actual temperature, we determined all the peaks that have occurred in that particular country. The predicted temperature at which the COVID-19 outbreak will peak was computed using equation 2 with central values of A1, B1, and C1. The prediction error was computed by taking the difference between the actual value and the predicted value. We could reach a prediction accuracy of up to 99 % and only in the case of New Zealand, the prediction accuracy was 41.4 %. in most of the cases the prediction accuracy was above 70 % in the case of New Zealand the predicted temperature peak did not match with the actual peak. the reason could be only one peak, and low population density.

**Table 1.** Results of temperature prediction with blind testing for 7 countries and corresponding accuracy on prediction

| Country Name | Latitude | Temperature at peak (actual) | Temperature at peak (predicted) | Error | Prediction Accuracy |
| --- | --- | --- | --- | --- | --- |
| Argentina | -38.4 | 13 | 9.10 | 3.90 | 0.700 |
| Kuwait | 29.3 | 19.6 | 20.63 | -1.03 | 0.950 |
| New Zealand | -40.9 | 16 | 6.62 | 9.38 | 0.414 |
| France | 46.2 | 8.6 | 9.98 | -1.38 | 0.861 |
| Italy | 41.9 | 11 | 13.03 | -2.03 | 0.844 |
| Indonesia | -0.8 | 27 | 26.72 | 0.28 | 0.990 |
| Egypt | 26.8 | 20 | 21.85 | -1.85 | 0.916 |

## 4.1 Discussions

We have observed that not all COVID-19 peaks can follow the latitude-based equation that we have developed. But on the other hand, many countries might have multiple series of covid-19 outbreak waves, out of these one may overlap with our prediction. There were typically 4 waves in most of the countries in a period of about 1.5 years. Due to vaccination, the COVID-19 virus will take some time to adapt itself and become a variant. This might lead to skipping an entire year from the COVID-19 outbreak. Though, we found that there is no straight correlation between temperature and COVID-19 outbreak. But there exists a slight possibility that along with latitude and temperature we can predict at what month of the year probability of COVID-19 outbreak is maximum. With this prediction in hand, Governments can plan their medical resources in advance for these months and avoid many deaths that might occur due to a shortage of these resources. This study was only limited to latitude and temperature with just numbers provided by different governments. So any kind of tampering to these COVID-19 numbers will badly affect our analysis and the entire prediction might fail in that case. We have not at all considered important parameters, such as population density, humidity, social distancing, and use of a mask by the people, availability, and awareness of vaccines. Though the variant of the COVID-19 virus can totally restart the pandemic the availability of its temperature-latitude dependant help us retune our equation and we can be prepared for the future. This study was based on the data availability of different countries and just a course prediction as we know not an entire nation can have the same temperature neither latitude. For larger counties like the USA and Australia, researchers can do similar studies statewide or district-wise also.

## 5 Conclusions

We have successfully designed an equation that can predict the temperature at which the COVID-19 outbreak will peak for the known latitude. We have developed the equation using the Levenberg–Marquardt algorithm with the help of the first-order sum of the sine equation. We have collected temperature data from 12 different countries and then blind-tested the data on 7 different counties. We found that 6 out of 7 countries did follow our prediction with more than 70 % accuracy. The maximum accuracy that we could achieve is 99 % and the average accuracy of prediction is around 81.1 %. We expect after knowing the temperature range each country can find its own days on which COVID-19 wave will occur. Also, we found bi-modal excitation of COVID-19, i.e. the COVID-19 outbreak is seen in lower temperature (0 *°*C to 13 *°*C) ranges as well as in the higher temperature ranges (21 *°*C to 35 *°*C). We expect the work will be carried forward by other researchers and may find more interesting results based on our analysis and eventually help society.

## Data Availability

Data will be made available in the public domain after the publication of this manuscript.

## Acknowledgements

The authors would like to thank all the colleagues of the A. P. Shah Institute of Technology.

## Declarations

### Code availability

Code will be made available in the public domain after the publication of this manuscript.

### Authors’ contributions

Conceptualization was done by P. P. Adivarekar (PA) and S. S. Aloni (SA). All the literature reading and data gathering were performed by M. A. Jain(MJ). The formal analysis was performed by PA, SA, and MJ. Coding was done by MJ and SA. Manuscript writing-original draft preparation was done by SA and MJ. Review and editing was done by PA. Visualization work was carried out by PA and SA.

### Ethics approval

All authors consciously assure that the manuscript fulfills the following statements: 1) This material is the authors’ own original work, which has not been previously published elsewhere. 2) The paper is not currently being considered for publication elsewhere. 3) The paper reflects the author’s own research and analysis in a truthful and complete manner. 4) The paper properly credits the meaningful contributions of co-authors and co-researchers. 5) The results are appropriately placed in the context of prior and existing research.

### Consent to participate

This article does not contain any studies with animals or humans performed by any of the authors. Informed consent was not required as there were no human participants. All the necessary permissions were obtained from the Institute Ethical Committee and concerned authorities.

### Consent for publication

Authors have taken all the necessary consents for publication from participants wherever required.

